# Improving neglected tropical disease services and integration into primary healthcare in Southern Nations, Nationalities and People’s Region (SNNPR), Ethiopia: results from a mixed methods intervention evaluation

**DOI:** 10.1101/2023.10.11.23296918

**Authors:** Laura Donovan, Tedila Habte, Esey Batisso, Dawit Getachew, Ann-Sophie Stratil, Agonafer Tekalegne, Fikre Seife, Damen Mariam, Kevin Baker

## Abstract

**Background:** Ethiopia is one of the countries with the highest burden of neglected tropical diseases (NTDs), with 16 of 20 recognised NTDs considered a public health problem, twelve of which have been identified as public health priorities by the Ethiopian Federal Ministry of Health. However, until recently NTDs have not received adequate attention at national and subnational levels in the country.

**Methods:** This study was conducted in Damot Gale district, Wolaita Zone in (Southern Nation Nationalities Peoples Region) SNNPR, Ethiopia and used a mixed methods approach to evaluate an intervention integrating four common NTDs (trachoma, lymphatic filariasis, schistosomiasis and podoconiosis) into Ethiopia’s primary healthcare system. The intervention consisted of adapted job aids, supportive supervision, and improved supplies of medical tools to improve diagnosis, management and reporting.

**Results:** found that the intervention was successful at improving the detection, management and reporting across the four common NTD’s included and had a high level of acceptance from health workers. The intervention demonstrated cost-effectiveness.

**Conclusion:** The findings highlight the need for further investment and consideration of integrating and scaling up NTD interventions at the primary healthcare level in Ethiopia, demonstrating that providing a package of interventions to support integration can be a cost-effective method.

**Author Summary:** Building on the findings of a previous small-scale operational study and formative phase, this study involved implementing an intervention to integrate the prevention, diagnosis, management and reporting of four common NTDs — trachoma, lymphatic filariasis, schistosomiasis and podoconiosis — into Ethiopia’s primary healthcare system. The intervention consisted of providing health workers with adapted job aids, supportive supervision and improved diagnostic and medical supplies to facilitate NTD diagnosis, management, and reporting. It was implemented for six months in one hospital, one health centre and five health posts in Damot Gale district, Ethiopia and the feasibility, acceptability and cost-effectiveness were evaluated. Results indicate that the capacity of all enrolled health facilities for detecting, managing, and recording target NTDs improved over time. The use of intervention materials by health workers also increased over time. The intervention tools proved to be highly acceptable to health workers who viewed them as helpful, relevant, and easy to use. The findings highlight that providing a package of interventions to support integration can be a cost-effective method and that the integration and scale of NTD interventions at the primary healthcare level in Ethiopia should be considered.

## Introduction

Neglected tropical diseases (NTDs) are a diverse group of diseases that are mainly prevalent in tropical areas, with over 40% of the global NTD burden concentrated in sub-Saharan Africa [1]. NTDs affect more than 1.5 billion of the world’s population, particularly communities with higher poverty rates, and women and children [1]. In recent years, the global development community has come together to tackle the burden of NTDs, agreeing on an ambitious NTD roadmap [2]. Reducing NTD morbidity and mortality is formally recognised as a target for global action in SDG 3.3, which calls to ‘end the epidemics of […] neglected tropical diseases’ to ‘ensure healthy lives and well-being for all at all ages’ [3]. Although a great deal has been achieved by NTD programmes during the past decade, progress towards achieving NTD and universal health coverage (UHC) targets will depend on bringing NTD programmes, functions and activities into the mainstream of broader health systems. In addition, efforts to innovate and intensify the management of NTDs must focus on ensuring that these diseases are detected and managed within primary healthcare systems [1]. The 2030 NTD global roadmap also reaffirms the importance of integrating efforts across NTDs and within national health systems in the context of UHC and coordinating stakeholders in NTDs and related areas [4].

Ethiopia is one of the countries with the highest burden of NTDs, with 16 of 20 recognised NTDs considered a public health problem, twelve of which have been identified as public health priorities by the Ethiopian Federal Ministry of Health [5]. However, NTDs have not received adequate attention at national and subnational levels in the country until recently. The Ethiopian Third National NTD Strategic Plan (2021-2025) lists the mainstreaming of NTDs into the routine healthcare delivery system as a cross-cutting and strategic priority [6]. In addition, nationwide NTD mapping has been conducted, control interventions for the various NTDs – including mass drug administration – have been scaled up, and a post-treatment surveillance system and strong NTD research forum have been established [7].

This study evaluated an intervention which integrates NTDs into primary healthcare. The intervention focused on strengthening the detection, management, recording and reporting of four NTDs; trachoma, lymphatic filariasis, schistosomiasis and podoconiosis. These NTDs were selected due to being identified as public health priorities in Ethiopia [5]. Although these NTDs differ in their transmission and management, there is overlap in the populations they affect, and the presentation of these diseases can lead to stigma [8][9][10]. Nationwide mapping exercises conducted between 2013 and 2017 revealed 412 national districts and 70 districts in (Southern Nation Nationalities Peoples Region) SNNPR were endemic for schistosomiasis [11]. Preventative chemotherapy, prompt diagnosis and treatment of schistosomiasis, as well as access to clean water and good hygiene practices, can help to reduce transmission. Ethiopia is also the most trachoma-endemic country in sub-Saharan Africa, with a prevalence of active trachoma of 25.9% and blinding trachoma of 1.5% in the Southern Nations Nationalities and Peoples’ Region (SNNPR) in 2016 [12]. Trachoma can be treated with antibiotics but in the case of repeated infection and if left untreated the disease can lead to blindness. Prompt treatment of trachoma and good hygiene practices can help to reduce transmission. Both lymphatic filariasis, caused by infection with filarial parasites, and podoconiosis, a non-infectious disease, can lead to the abnormal enlargement of body parts, causing pain and severe disability. Nationwide mapping exercises conducted in 2017 revealed 71 national-level districts and 24 regional-level districts were lymphatic filariasis-endemic [13]. Lymphatic filariasis preventative treatments are used to prevent infections but once the disease has progressed management of swelling in affected areas is the only option. As of 2021, the average prevalence of podoconiosis in Ethiopia was 4% (an estimated 1.56 million cases), with the highest prevalence in SNNPR (8.3%) [14]. Treatment for this disease is limited to lymphoedema management. For each of these NTDs management can help to alleviate suffering and reduce stigma associated with the presentation of some of these diseases.

A previous small-scale operational research study was conducted in 2018 and provided the basis for this larger study, conducted between 2019 and 2021 [15]. This study comprised of two phases a formative phase and an intervention phase. The formative phase identified gaps in the integration of NTD services in the Ethiopian primary healthcare system and was used to inform the development of a tailored, multi-component intervention. Data collected during the formative phase provided a baseline for evaluating the health system capacity component. The intervention phase piloted the intervention in selected health facilities for six months and evaluated for feasibility, acceptability, and implementation cost. This paper presents the results of this evaluation and outlines recommendations to ensure the sustainability of future NTD service integration within the Ethiopian primary healthcare system.

## Methods

### Study setting

The study was conducted in Damot Gale district, Wolaita Zone in SNNPR, Ethiopia. This district was selected due to the endemicity of the four target NTDs and operational feasibility due to the proximity to Malaria Consortium’s sub-country office in Hawassa. Damot Gale is located 350 km south of Ethiopia’s capital, Addis Ababa, and 153 km southwest of Hawassa. The district is mainly rural and is subdivided into 31 *kebeles*, with a population of approximately 183,720 as of 2017 [14][16]. Within Damot Gale, Boditi primary hospital, Buge health centre and five rural health posts were selected to implement the intervention. Inclusion criteria included having an adequate health workforce, access to electricity, clean water and sanitation, no enrolment in other research studies and not receiving support from non-governmental organisations.

### Study design

The aim of this study was to evaluate the package of interventions developed to support the integration of four common NTDs into the health system. The package of interventions included: clearly outlining staff roles and responsibilities, providing clear and harmonized NTD case definitions, providing job aids for integrated NTD detection and management, introducing sensitive diagnostic tests for intestinal schistosomiasis, training for health workers and Health Extension Workers (HEWs), provision of drugs and medical supplies, inclusion of NTD supervision tools, indicators for suspected targeted NTDs and social and behaviour change communication (SBCC) materials. The intervention was evaluated using a mixed methods approach to determine the feasibility, acceptability and cost-effectiveness. The evaluation comprised of:

- Two observational assessments; one immediately after the health worker training, and one after the six-month intervention pilot. These aimed to determine the proportion change in NTD assessments demonstrating adequate knowledge of target NTDs by health workers and their use of the intervention materials.
- A health system capacity assessment to evaluate changes in the capacity of the study health facilities to detect, manage, record and report target NTDs.
- The collection and analysis of routine monitoring and evaluation (M&E) data from the study health facilities to identify any changes in NTD prevalence and service delivery post-intervention.
- Key informant interviews (KIIs) with health workers, HEWs, health development army members (HDAs) and community members to elicit their perceptions of the intervention’s reach, fidelity, acceptability and feasibility.
- A cost-effectiveness analysis to determine the intervention’s value for money.

### Data collection and analysis

Baseline data were collected during the formative phase and in April 2021 immediately after the health worker training. Endline data were collected in October 2021 six-months after the start of the intervention. Data collection procedures and data collector training were compliant with national COVID-19 prevention guidelines, with all participants wearing a facemask, maintaining physical distancing, washing hands frequently and regularly disinfecting materials. All data collectors had a first degree or higher, experience within the Ethiopian healthcare system and spoke Amharic, English and Wolaytegna.

#### Observational assessments

43 patients with signs and symptoms of target NTDs were enrolled and participated in the baseline observation assessment in April 2021. In the endline observation assessment, conducted in October 2021, a total of 129 patients with signs and symptoms of target NTDs were enrolled and participated. Data for the observational assessments was collected by a trained research assistant observing health workers during patient consultations. To assure data quality, data collectors participated in a three-day training beforehand which included a pre-test of the observation checklists. During each consultation the research assistant observed and recorded data using a checklist developed for each facility type. The research assistant recorded the signs and symptoms the patient presented with, patient history, the type of diagnosis or test (clinical examination, laboratory test) received, the health worker’s diagnosis, the health worker’s recommendations in terms of disease management (treatment, follow-up, home management), the treatment prescribed, if the patient was referred, if the health worker recorded the diagnosis and the services provided, and if the health worker consulted any guidelines or reference documents during the consultation.

Baseline and endline observational assessment data were quantitatively analysed using SPSS to determine the changes in health workers’ knowledge during patient consultations, adherence to intervention procedures, health worker ability to detect NTDs and the acceptability of the intervention tools.

#### Health system capacity assessment

The health system capacity assessments were conducted by two trained data collectors. Structured questionnaires were used which covered the health facility profile and the following areas related to NTDs: general infrastructure, supervision, drugs and equipment, laboratory infrastructure and equipment, guidelines and job aids, health personnel, training and recording and reporting. Two questionnaires were used: one for Boditi hospital and Buge health centre and one for the health posts. Health posts do not have a laboratory and so questions on laboratory infrastructure and equipment were omitted. The questionnaires were input into the data collection app CommCare and data were collected electronically using tablets. To assure data quality, data collectors’ skills were evaluated during a pilot prior to data collection, data submitted in CommCare were checked daily and standard operating procedures (SOPs) were developed to ensure consistency in data entry. Health system capacity assessment data were exported from CommCare into Excel for analysis. The data were analysed quantitatively and qualitatively based on the designed relevant six thematic areas: general infrastructure, supervision, drugs and equipment, laboratory infrastructure and equipment, guidelines and job aids, and training.

#### Routine M&E data

Routine NTD M&E data were retrieved from health facility records, which are captured using two separate forms: one for diseases and one for service delivery, both through a DHIS2 tool. As HEWs are not tasked with NTD reporting, reporting forms were prepared by the research team for HEWs to report suspected target NTDs. Results from schistosomiasis-related laboratory tests were also recorded before and after the introduction of the intervention. Routine M&E data were analysed quantitatively by inputting the collected data into Excel. The analysis was done based on data collected on laboratory tests, disease reports related to target NTDs (DHIS2), service delivery reports related to target NTDs (DHIS2) and a parallel data reporting form developed by the research team.

#### KIIs

KIIs were conducted in October 2021 with 10 HEWs from five health posts (2 per health post), 8 health workers (4 from health centre and 4 from hospital level), 4 laboratory technicians (2 from health centre and 2 from hospital level), 2 data managers (1 from health centre and 1 from hospital level) and 5 HDA members. All participants were involved in either the detection, management or recording and reporting of NTDs at their health facility and were exposed to the intervention during implementation. KIIs were also held with male and female community members; 6 people living with disability caused by a target NTD and 6 living without disability. Topic guides were designed to explore participant perceptions of the reach, fidelity, acceptability and feasibility of the intervention, including their knowledge of the four target NTDs and their control. Health worker KIIs took place at their respective health facility and community member KIIs were held in a neutral space such as a school. All KIIs were conducted by trained data collectors. To ensure data quality, all data collection procedures and tools were piloted for one day, with separate pilots for each local language. KIIs were conducted in the participant’s preferred language (Amharic, English or Wolategna) and audio recorded using a digital recorder.

Community-level key informants were identified and selected through convenience sampling, based on their availability and willingness to participate. Health system-level informants were identified and selected by the Malaria Consortium team based in Hawassa on the basis of their role and experience of working on NTDs within the Ethiopian primary healthcare system.

A thematic analysis was conducted for data collected during KIIs [17]. Audio recordings were transcribed verbatim and translated into English where applicable. For those KIIs not conducted in English, data quality and translation accuracy were assured by checking a sample of the transcripts and backtranslating these into the local language. Transcripts were read to generate a coding list, which was then applied to all transcripts. The software MAXQDA 2020 was used to manage, code and retrieve data. Potential themes were identified and data relevant to each theme were collated; these were then discussed among the research team before final consolidation. Theme names were agreed by the research team and each theme description was refined and substantiated with compelling participant quotes.

#### Cost-effectiveness

A cost-effectiveness analysis was conducted to measure the resource flow or total value of resources used to improve NTD services through integration into primary healthcare. A cost-effectiveness analysis compares an intervention’s costs to its outcomes. It expresses outcomes in natural health units, in this case the proportion change in terms of primary healthcare capacity in detecting and managing target NTDs through the introduction of the intervention, as well as the system’s capacity to produce relevant, timely and accurate data with regard to disease burden. The ratio of net programmatic costs vs. net programmatic effects was determined and the costs incurred per disability-adjusted life year (DALY) averted with that of the current gross domestic product (GDP) per capita compared.

The TreeAge Pro 2021 software was used for the cost-effectiveness analysis. The software constructs a decision analytic model with a Markov process to estimate the programmatic costs incurred and the health impacts (in terms of DALYs averted) within the project. The decision tree model follows a series of steps to construct a tree structure under uncertainty for alternative options and selects the least expected cost per effect as the best alternative among the constructed steps.

### Informed consent and ethical considerations

Prior to the observation assessments and KIIs, participants were given an information sheet and consent form to read which included information on the withdrawal of consent at any point and the contact details of the study coordinator. Participants were allowed up to an hour to confirm their participation and were guided through the information sheet to assist their decision. For illiterate participants, data collectors read the information sheet and consent form aloud in the local language and confirmed their consent to participate by marking their digital fingerprint.

To ensure participant confidentiality and data authenticity, audio files and transcriptions of KIIs were kept in secure, locked cabinets, accessible by the Malaria Consortium research study team. Paper consent forms and KII topic guides were stored in opaque carriers at all times. A unique identification code was used to anonymise subject data (e.g. 1001) and subject identifiers were not used on data forms or during KIIs. Data stored in electronic formats were password protected.

This study was approved in June 2020 by the SNNPR Public Health Institute Research Ethical Committee. However, implementation of study activities was delayed due to COVID-19-related restrictions imposed by the Ethiopian government. To address these delays, an amendment agreement between The Task Force for Global Health and Malaria Consortium was signed for a costed extension of the project by nine months until December 2021.

## Results

### Observational assessment of service providers

43 patients with signs and symptoms of the four target NTDs were enrolled and participated in the baseline observation assessment. In the endline observation assessment, a total of 129 patients with signs and symptoms of target NTDs were enrolled and participated. The participant characteristics are summarised in supplementary information Table 1.

**Table 1.** Proportion of assessment stages conducted at baseline and endline.

| Assessment stage | Baseline (%) | Endline (%) | Percentage point difference (%) |
| --- | --- | --- | --- |
| Appropriate consideration of NTDs | 29/43 (67.4) | 114/129 (88.4) | 21.0 |
| Consultation of job aid during assessment | 26/43 (60.5) | 108/129(83.7) | 23.2 |
| Use of appropriate job aid for the age group | 26/43 (60.5) | 104/129 (80.6) | 20.1 |
| Linking of iCCM/IMNCI guidelines with NTD job aid for children under five | 1/4 (25.0) | 14/18 (77.8) | 52.8 |
| Appropriate application of examination procedures | 36/43 (83.7) | 107/129 (82.9) | -0.8 |

#### Adherence to intervention procedures

The proportion of each of the five assessment stages (outlined in table 1) was used to calculate the proportion of NTD assessments demonstrating adherence to intervention procedures by health workers at baseline compared to endline. At endline, proportion change improvement, which ranged from 20% to 53%, was shown in four out of the five assessment stages, compared to the baseline. The proportion of one assessment stage, ‘appropriate application of examination procedures’, showed a minor decrement (-1%) at endline compared to baseline.

### Health system capacity assessment

Table 2 summarises the results of the health system capacity assessment conducted before and after the intervention pilot at Boditi primary hospital, Buge health centre and the five rural health posts. Overall, there was a marked increase in the capacity of the primary healthcare system to detect, manage and record NTDs post-intervention implementation. More attention was given to the integration of NTDs into routine services, which previously only focussed on mass drug administration (MDA).

**Table 2.** Results of health system capacity assessment pre-and post-intervention implementation.

| Assessment area | Pre-intervention | Post-intervention |
| --- | --- | --- |
| General infrastructure | *No dedicated triage room for NTDs<br>*Trichiasis surgery not available | *Dedicated triage room created<br>*Trichiasis surgery available and 5 surgeries performed |
| Supervision | *†# Supervision checklists did not address target NTDs | *†# Supervision checklists developed by the study that address target NTDs routinely used |
| Drugs and equipment | *†# No drugs or equipment available for treatment of target NTDs and management of lymphedema | *† Some drugs and equipment for the treatment of target NTDs and management of lymphedema available, as provided by the study<br>Lymphedema management initiated (*) and intensified (†) |
| Laboratory infrastructure and equipment (* and † only) | *† No circulating cathodic antigen (CCA) diagnostic tests available for schistosomiasis (only wet mount microscopy) | *† CCA tests made available by the study and routinely used, with an increase in detected cases recorded |
| Guidelines and job aids | *†# No job aids and case definitions on target NTDs available | *†# Job aids on target NTD detection, management and recording and case definitions available and routinely used, as developed by the study<br>*†# Guidelines for CCA tests available |
| Training | *Only 2 doctors attended training on lymphedema management and prevention<br>† Only 1 health officer and 1 clinical nurse attended trichiasis surgery training<br># Most HEWs trained only on trachoma clinical diagnosis and treatment | 40 health personnel (*), 29 health personnel (†) and 9 HEWs (#) attended comprehensive training on target NTDs, as organised and delivered by the study team |

\*Boditi hospital, † Buge health centre, #Health post

### Routine M&E data collection and analysis

Retrieved routine laboratory data related only to intestinal schistosomiasis as the intervention assigned point-of-care circulating cathodic antigen (CCA) diagnostic tests for this NTD at the hospital and health centre level. Prior to the intervention pilot, which took place from July to September 2021, these health facilities used microscopy for schistosomiasis detection in wet mount stool samples. In Boditi hospital, before the intervention pilot, of the total 942 stool samples taken with wet mount microscopy, no schistosomiasis cases were detected, possibly due to the reduced sensitivity compared to point-of-care CCA tests or microscopic quality issues. After introduction of the intervention, 16 tests were performed using CCA test kits, three of which were positive. As a result of the intervention the case detection rate increased from 0% to 18.75%.

In Buge health centre, before the intervention pilot, a total of 83 stool examinations were performed using microscopy, with only one case positive for schistosomiasis. After the intervention pilot a total of 181 CCA tests were performed, of which 65 schistosomiasis cases were detected. As a result of the intervention the case detection rate increased from 1.2% to 35.91%. These results are summarised in Table 3.

**Table 3.** Schistosomiasis case positivity rate before and after intervention implementation.

| Health facility | Baseline (Microscopy) |  |  | Endline (CCA) |  |  |
| --- | --- | --- | --- | --- | --- | --- |
|  | Tested | Cases | Positivity rate (%) | Tested | Cases | Positivity rate (%) |
| Boditi Hospital | 942 | 0 | 0.00 | 16 | 3 | 18.75 |
| Buge Health Centre | 83 | 1 | 1.20 | 181 | 65 | 35.91 |
| Total | 1025 | 1 | 0.09 | 197 | 68 | 34.52 |

#### DHIS2 disease reports

Prior to the intervention, target NTDs were not reported in DHIS2 reporting forms at Boditi hospital or Buge health centre. However, after the intervention, 3 schistosomiasis cases and 3 trachoma cases at Boditi hospital and 51 schistosomiasis cases at Buge health centre were reported. No lymphatic filariasis or podoconiosis cases were recorded during the reporting period at either health facility. However, this is because these diseases are not included in the DHIS2 disease reporting tool.

#### DHIS2 service delivery reports

Results of NTD service delivery data, which are reported electronically through DHIS2 on a quarterly basis, are presented in Table 4. At Boditi hospital, there were no reported NTD services before the intervention, however lymphedema management and trichiasis surgery were initiated after the six-month pilot period. At Buge health centre, the lymphedema management and trichiasis surgery were intensified after the intervention period, with the number of lymphedema cases increasing fivefold. There were no hydrocele operations reported at Boditi hospital or Buge health centre before or after the intervention.

**Table 4.** NTD service delivery before and after intervention implementation.

| Health facility | Baseline |  |  | Endline |  |  |
| --- | --- | --- | --- | --- | --- | --- |
|  | Lymphedema management | Trichiasis surgeries | Hydrocele operations | Lymphedema management | Trichiasis surgeries | Hydrocele operations |
| Boditi hospital | 0 | 0 | 0 | 12 | 5 | 0 |
| Buge health centre | 49 | 26 | 0 | 283 | 38 | 0 |
| Total | 49 | 26 | 0 | 295 | 43 | 0 |

#### Health post data

There is no formal system for recording and reporting disease data at the health post level, therefore monthly parallel reporting forms were developed as part of the intervention. For this reason no suspected target NTDs were recorded before the intervention period. After the provision of parallel reporting forms and training on their use, all health posts initiated recording suspected NTD cases and referral to the nearest health centre. The suspected cases recorded before and after the intervention implementation are shown in Table 5.

**Table 5.**
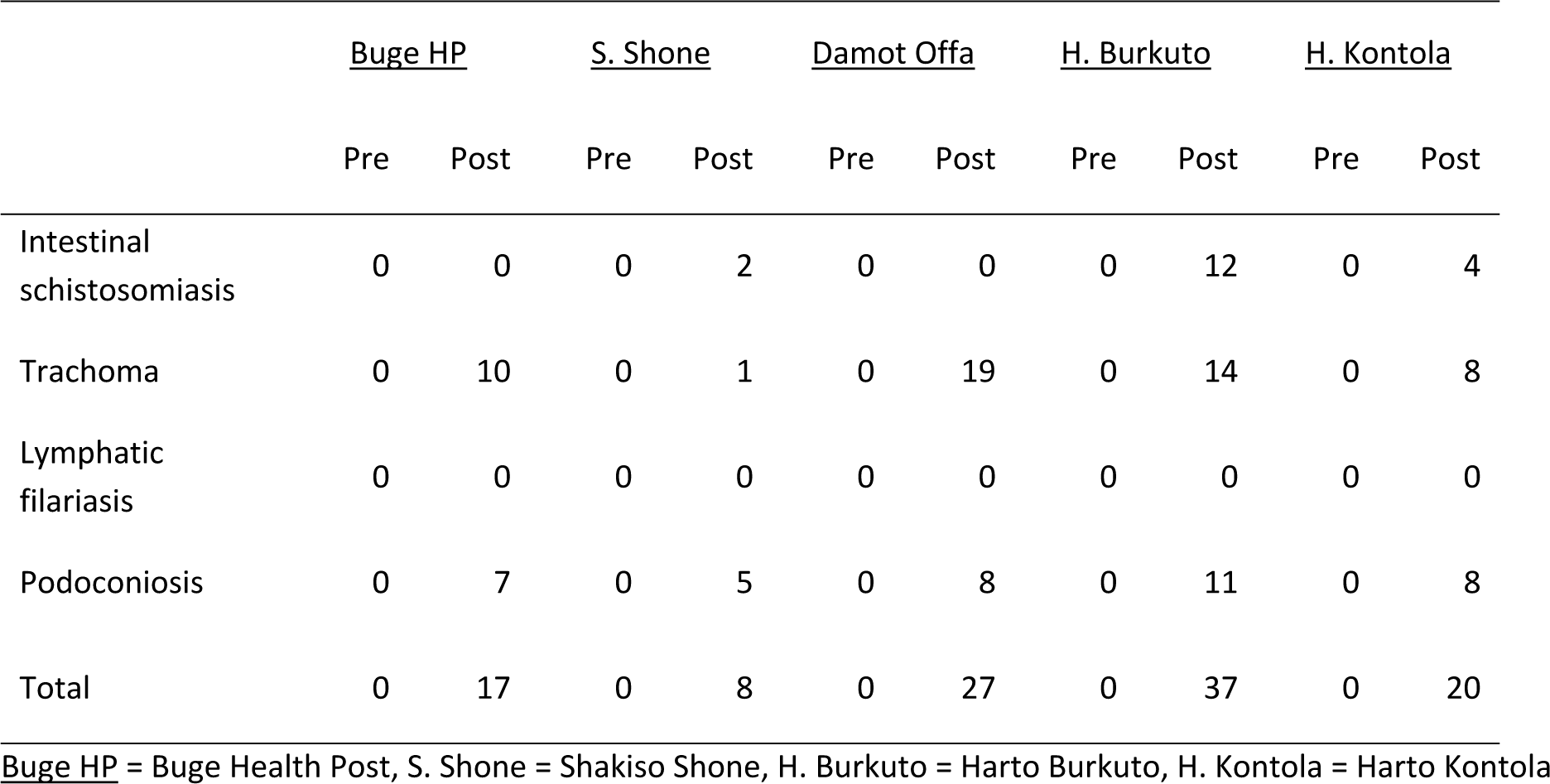
suspected NTD cases reported at health posts before and after intervention implementation.

### Key informant interviews

The following results are from KIIs held with health staff and community members. Health staff included those at the health post, health centre and hospital level who were involved in either the detection, management or reporting of NTDs at their health facility and were exposed to the intervention during implementation. Community members included both those living with and without disability caused by the target NTDs.

#### Acceptability and usability of training materials received by health staff

Most participants received materials during the training, including guidelines on NTD detection, management and follow-up, manuals, registration books, referral forms and job aids. Materials were used regularly to assist with NTD detection and management, to advise other staff and to determine the appropriate drug dosage. Participants described the materials as clear, easy to understand, user-friendly and informative. Most agreed that the materials contained a sufficient amount of information and adequately considered non-NTDs (e.g. malaria), aided accurate NTD diagnosis, and reduced the risk of under-or over-diagnosis. Suggestions for improvement were the inclusion of targets (e.g. how much follow-up a case should have), possible complications and other diseases with overlapping symptoms to aid differential diagnosis. Many expressed that training should be given continuously to ensure the sustainability of learnings and regular review meetings should be held to assess the impact on daily activities.

> *“I was using this manual to remember what I learned and in the case of detection as guiding tools, also as a reference to manage the cases most of the time podoconiosis.”* (Health worker, Buge health centre)
>
> *“They are also easy to use. They came in Amharic, they are easy for us. The content is good.”* (HEW, Buge rural health post)
>
> *“It should be given in sustainable manner, it should not be done in campaign for a week… there should be continuous follow up after the end of training because one time event may not last long.”* (Health worker, Boditi Hospital)

#### Health staff and community member perceptions of tasking health workers with NTD management

Overall perceptions were positive, with most believing health workers have sufficient capacity to manage NTDs. It was felt that as they are present in the communities and many individuals are reluctant to seek care at higher health facilities due to financial and physical barriers, this will improve access to healthcare. Challenges raised mainly related to additional workload and a lack of resources such as drugs, equipment and staff. Future recommendations included training healthcare staff to address knowledge gaps, conducting community outreach activities to access patients in rural areas and strengthening collaboration with key stakeholders to raise community awareness of NTDs.

> *“This work put an additional burden on health workers at all levels, in my point of view we were performing very well.”* (Health worker, Buge health centre)
>
> *“To do this work it needs community engagement, strong integration or linkage with health post, health centre and hospital and support from woreda and NGO.”* (HEW, Shakiso Shone health post)

Community member perceptions on tasking HEWs with NTD control were mixed. For some, “it is a very good thing”, they should be encouraged to do so and “we need to support them to identify those cases and bring them to health facilities”. Others did not foresee any challenges and believed that as people are aware of the benefits of seeking care and being educated on NTDs, they will be willing to listen to and cooperate with the HDA and HEWs. Although supportive of the idea, some participants mentioned the need to ensure all the necessary resources are available beforehand, such as drugs and equipment. However, a few community members had negative perceptions, describing how NTD-related problems are beyond the capacity of the HDA and HEWs and “they can’t treat such problems”. For a community member with disability, the additional task would not have a positive impact on health outcomes as drugs will still remain unaffordable for many. A CMWO believed that the HDA are not able to “differentiate and diagnose people who develop disability”.

> *“Problems like mine are beyond their capacity. They can’t do anything about that. It is beyond what they can do.”* (Male community member with disability)
>
> *“Both health extension workers and the HDA can manage NTD cases if the equipment is available.”*
>
> (Female community member without disability)

#### Health staff perceptions of the intervention

The vast majority of health workers felt confident to perform NTD responsibilities during the intervention, mainly due to the training and materials received. However, a small proportion of HEWs did not feel fully confident due to the training being “too short”. Most participants also believed in the intervention’s approach for ensuring reliable NTD detection and noticed an improvement in patient outcomes. The main challenge spanning all four NTDs was the shortage of drugs and equipment to perform tasks. The impact on workload was also raised, except for schistosomiasis, which resulted in many health staff feeling this may affect the sustainability of the intervention.

> *“I am ready to handle any case according to the training I took. Every professional should treat patients according to the training they took and everybody should treat.”* (Health worker, Buge health centre)
>
> *“There is workload due to the human resource scarcity and the trained health professionals have double responsibility.”* (Health worker, Boditi Hospital)
>
> *“For the other problems it is better if we get training again. Also health workers as well as other health officers should support us.”* (HEW, Buge Rural health post)

#### Recommendations for NTD integration into primary healthcare

Health staff made several recommendations for integrating the detection, management, recording and reporting of NTDs into primary healthcare. These included having health workers and community leaders conduct awareness-creation activities within communities to “use every opportunity to deliver messages that can bring significant change”. Ensuring the necessary supplies (e.g. drugs and equipment) are available was frequently highlighted as an important factor to ensure the sustainability of NTD services. It was suggested that drugs and services should be provided free of charge as those susceptible to NTDs are often of low socioeconomic status, which may in turn encourage more healthcare-seeking. Having a dedicated focal person “who will work only on NTD-related issues at each level” to reduce the work burden on HEWs was also raised, this focal person should be incentivised to boost their motivation. Other recommendations included continuous support and follow-up on health workers from higher bodies and non-governmental organisations and regular trainings. This should also involve kebele leaders. Review meetings to evaluate these activities, including experience-sharing between different kebeles may facilitate future improvements in service provision. For one health worker, cooperation within the primary healthcare system and having a well-structured health post-hospital linkage was critical.

> *“Services should be given in a continuous manner as part of routine services to sustain the intervention results. The data which are being reported to higher bodies should also be evaluated like other services.”* (Health worker, Boditi Hospital)
>
> *“When the healthcare worker is taking history he has to check if there are any signs and symptoms related to NTDs and take action. This is how the services should be integrated.”* (Laboratory technician, Buge health centre)

### Cost-effectiveness analysis

#### Intervention cost

A summary of the average total costs is shown in Table 6. Accordingly, the cost of the intervention is summarised as: total capital (130,310 birr) and total recurrent (19,577,101 birr). The overall total adds up to 19,708,411 birr or USD 450,478 based on the exchange rate of 1 USD to 43.75 birr that prevailed in the first week of July 2021. Using a total beneficiary population of 302,842, the per-capita unit cost<colcnt=2>

**Table 6.** Total and unit costs of intervention implementation.

|  | Boditi hospital | Buge health centre |
| --- | --- | --- |
| Capital (birr) |  |  |
| Equipment | 9 660.00 | 16 650.00 |
| Transportation rental | 60 000.00 | 45 000.00 |
| Total capital costs | 69 660.00 | 61 650.00 |
| Recurrent (birr) |  |  |
| Personnel | 15 472 056.48 | 3 649 728.00 |
| Supplies | 115 180.12 | 157 916.40 |
| Vehicle operation and maintenance | 125 120.00 | 52 100.00 |
| Building operation and maintenance | 5 000.00 | - |
| Total recurrent costs | 15 717 356.60 | 3 859 744.40 |
| Total annual costs (birr) | 15 787 016.60 | 3 921 394.40 |
| Overall total annual costs (birr) | 19 708 411.00 |  |
| USD equivalent (1 USD = 43.75 birr) | 450 477.96 |  |

#### Outcome of integration

The main outcome measures at baseline and endline are shown in Table 7. The main outcome measures used for the effectiveness calculation included: the proportion of NTD assessments demonstrating adherence to target NTD intervention procedures by health workers (primary); the proportion of change in heath workers’ ability to detect target NTDs over time (secondary); the proportion of change in heath workers’ ability to detect and mange target NTDs over time (secondary); and the proportion of health workers and HEWs who can adequately detect and record target NTDs at the end of the intervention period (secondary). These measures are again converted to gains in years of life lived without disability (gains in YLL) based on the Global Burden of Diseases Study for Ethiopia for 2017 [18].

**Table 7.** Main outcome measures at baseline and endline.

| Indicator | Baseline (%) | Endline (%) | Percentage point difference (%) |
| --- | --- | --- | --- |
| NTD assessments demonstrating adherence to intervention procedures by health workers | 72.1 | 77.5 | 5.4 |
| Health workers able to detect target NTDs | 81.8 | 92.1 | 10.3 |
| Health workers able to detect and manage target NTDs | 81.8 | 81.2 | -0.6 |
| Health workers and HEWs able to detect and record target NTDs | 81.8 | 87.1 | 5.3 |
| Correctly diagnosed NTDs | 0.09 | 34.52 | 34.43 |

#### Cost-effectiveness of service integration

Assuming the effectiveness of the intervention (gains in health workers’ ability to detect NTDs) to be 34.43, the cost-effectiveness ratio of the intervention would be 21.23 (730.94/34.43). This ratio (730.94) results from dividing the total annual cost of intervention (USD 450,478) by the estimated decrease in DALYs lost (616.3) due to the increased ability of detection that came about following the integrated interventions. Based on comparison with the mean per-capita GDP prevailing at the period of project implementation (the willingness-to-pay threshold of one times the GDP per capita), the intervention can be considered cost-effective since the per capita cost of 730.94 USD is below the GDP per capita (USD 936.34) that prevailed during the intervention period. The input parameters for the cost-effectiveness model are outlined in Table 8. The cost-effectiveness model is shown in Figure 1.

**Figure 1.**
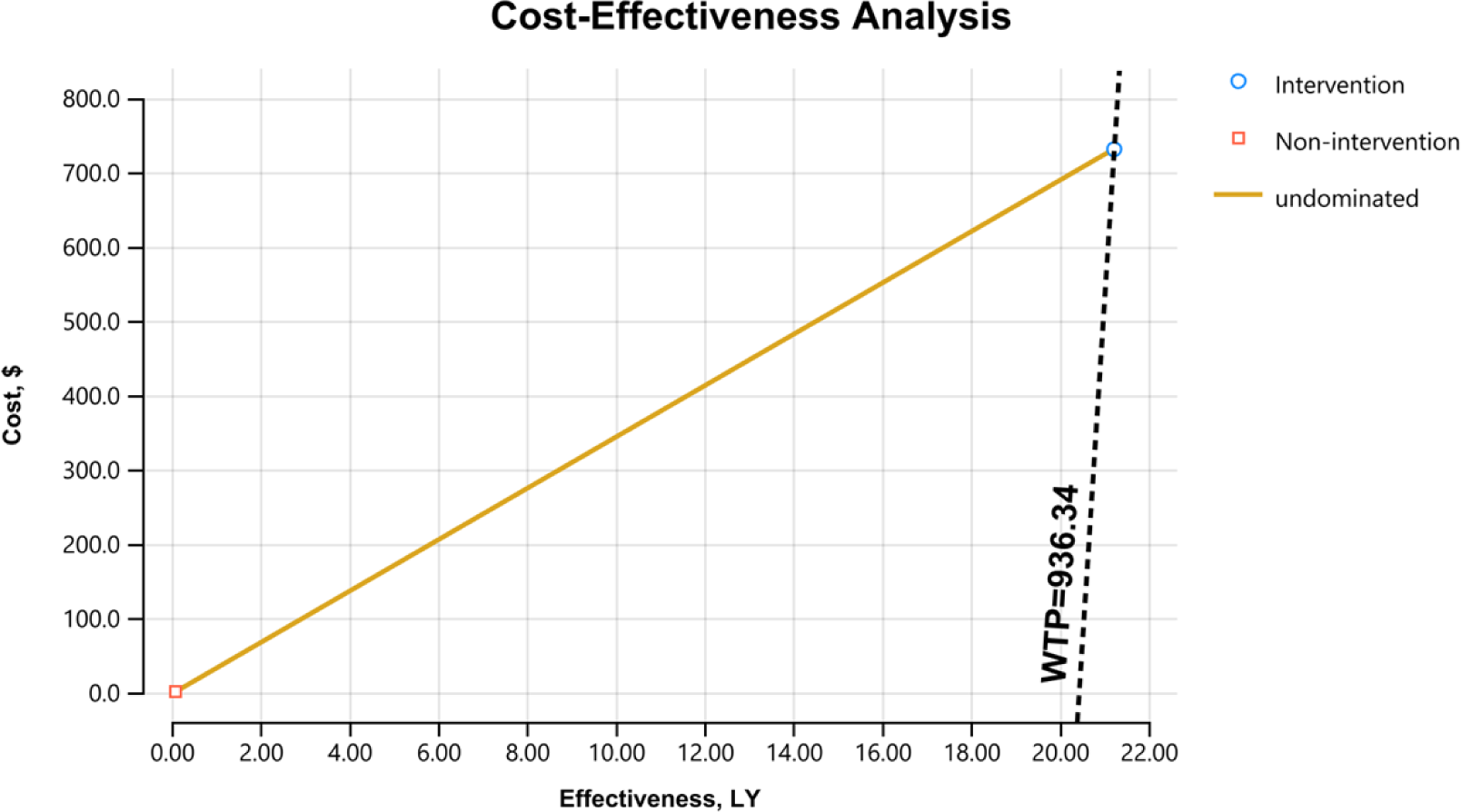
Cost-effectiveness model.

**Table 8.**
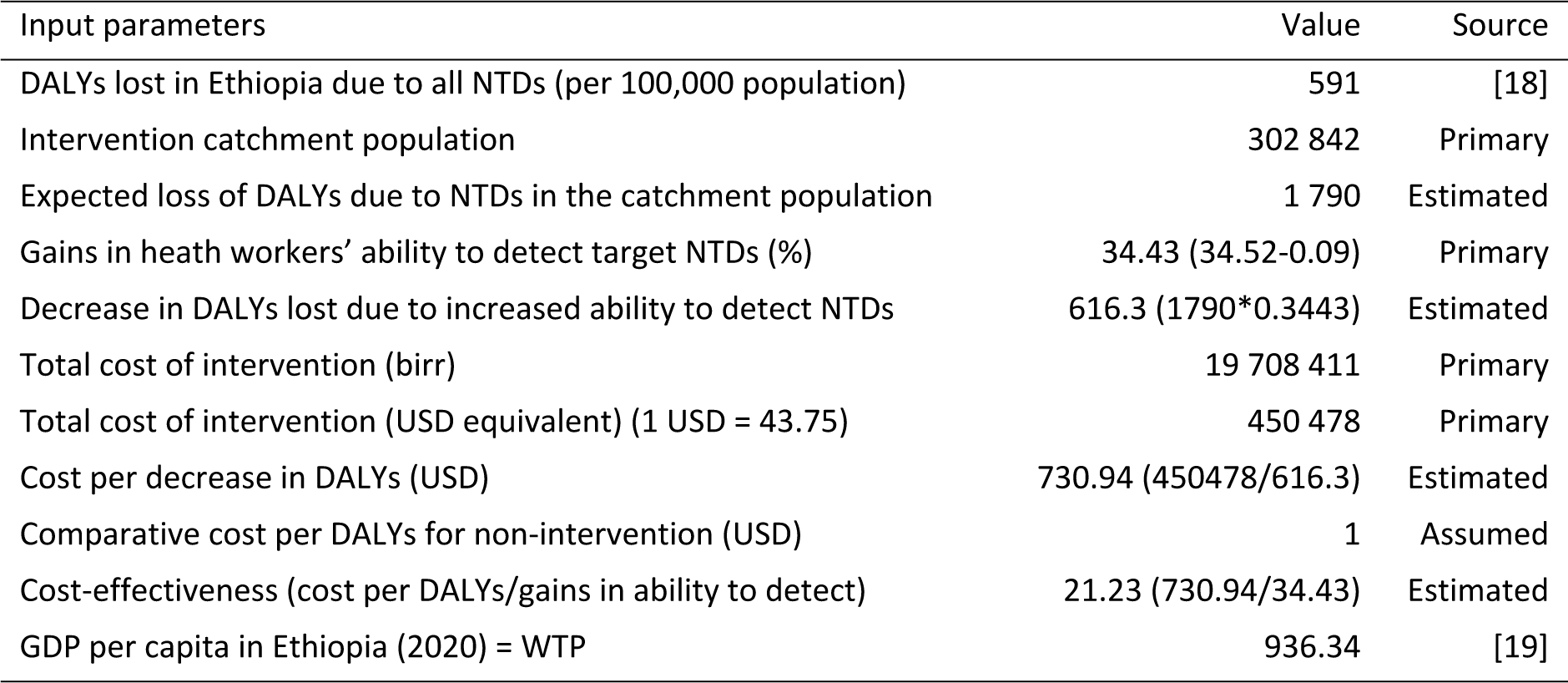
Input parameters for the cost-effectiveness model.

| Input parameters | Value | Source |
| --- | --- | --- |
| DALYs lost in Ethiopia due to all NTDs (per 100,000 population) | 591 | [18] |
| Intervention catchment population | 302 842 | Primary |
| Expected loss of DALYs due to NTDs in the catchment population | 1 790 | Estimated |
| Gains in health workers' ability to detect target NTDs (%) | 34.43 (34.52-0.09) | Primary |
| Decrease in DALYs lost due to increased ability to detect NTDs | 616.3 (1790*0.3443) | Estimated |
| Total cost of intervention (birr) | 19 708 411 | Primary |
| Total cost of intervention (USD equivalent) (1 USD = 43.75) | 450 478 | Primary |
| Cost per decrease in DALYs (USD) | 730.94 (450478/616.3) | Estimated |
| Comparative cost per DALYs for non-intervention (USD) | 1 | Assumed |
| Cost-effectiveness (cost per DALYs/gains in ability to detect) | 21.23 (730.94/34.43) | Estimated |
| GDP per capita in Ethiopia (2020) = WTP | 936.34 | [19] |

## Discussion

Overall, results from the study were promising. The health system capacity assessment revealed encouraging changes in the functions of the study hospital and health centers after the implementation of the intervention. Results showed that target NTD service delivery improved after the six-month intervention pilot, particularly in lymphoedema case management for lymphatic filariasis and podoconiosis, which can be attributed to the provision of training, job aids, drugs, supplies and materials. This reflects a similar result from the recent EnDPoINT study, also conducted in Ethiopia, which highlighted the need for continuous on-the-job training and supervision to maintain quality service delivery [13].

The study found there is a need for improved M&E data collection and analysis to support improved NTD case management. Overall, data on the target NTDs showed remarkable improvements post-intervention, which can be attributed to the positive impact of the intervention on service delivery. For example, there was a dramatic increase in the number of schistosomiasis cases detected at hospital and health centre level, from one case (with microscopy) pre-pilot intervention, to 68 cases through CCA tests during the intervention implementation, demonstrating the improved sensitivity of the tests. However, a significant disparity between laboratory data and data reported through DHIS2 was observed pre-and post-intervention at Buge health centre, which may be attributable to errors in recording and reporting. Furthermore, the number of trachoma cases reported through DHIS2 appeared relatively low, considering the intervention areas are trachoma-endemic. Further investigation for corrective action is needed to fully understand this.

Our study demonstrated increases in the reporting of suspected schistosomiasis and trachoma cases at health posts, however further study is needed to determine the intervention’s impact on the reporting of suspected lymphatic filariasis. In addition, further effort is needed to advocate for the integration of podoconiosis into the DHIS2 reporting platform. Given the importance of effective and efficient data reporting highlighted by the study, due attention is needed at Ministry of Health level to include NTD case reporting at the health post level, as this will enable the true burden of NTDs to be documented and the proper referral and management of cases tracked. This was further highlighted by a recent study conducted in Kenya, which also showed low-performance surveillance in relation to NTDs and the need to strengthen this going forward [20]. Similarly, work done by Sightsavers in Ethiopia calls for the improved use of surveillance and data integration to eliminate trachoma [21].

The study found improvements in the primary outcome, adherence to target NTD intervention procedures by health workers over time, with a 5.4 percentage point improvement between baseline and endline (from 72.1% to 77.5%). This was slightly lower than expected and could have been due to the influence of the pandemic on service delivery, with many health systems suffering from service delivery issues during this period [22]. However, when specifically observing the ability to detect target NTDs over time, a notable increase of over 10% was discovered. This could reflect the focus on diagnosis as part of the intervention and the provision of improved diagnostics, as well as job aids. These results reflect the findings from other recent work conducted to support NTD case management [23]. These results are further supported by looking at the associated actions over time, such as consulting job aids (increased from 60.5% to 83.7%) and the use of the appropriate job aid for the age group (increased from 60.5% to 80.6%).

The qualitative findings demonstrated widespread acceptability of the intervention by health workers and HEWs at the end of the intervention period, and participants reported increases in knowledge and skills, and positive perceptions of the job aids’ ease of use. Many health workers reported feeling confident managing NTDs since the training, however others said additional training would be beneficial. Health staff and community members also mentioned some persisting contextual barriers that could affect the effectiveness of the intervention. These barriers included: a lack of human resources, the availability of equipment and the affordability of treatment.

Considering ‘gains in health workers’ ability to detect target NTDs’ as a main outcome measure, there was a 34.43% percentage point increase. The difference (as measured in total DALYs averted) amounts to a gain of 616.3 life years among beneficiaries in the catchment area of the programme, and the cost-effectiveness ratio of the intervention is 21.23 (730.94/34.43). Therefore, as the per capita cost of the intervention (USD 730.94) is less than the mean per capita GDP prevailing during the intervention period (USD 936.34), the intervention can be considered cost-effective.

Overall, the findings from this study highlight the need for further investment and consideration of integrating and scaling up NTD interventions at the primary healthcare level in Ethiopia, and that providing a package of interventions to support integration can be a cost-effective method. Routine health services for morbidity management and disability prevention caused by NTDs are still lacking, even though they impose a huge burden on affected individuals and their communities in terms of physical and mental health, and psychosocial and economic outcomes. More must be done to improve the prevention, diagnosis, treatment and management for these diseases to move towards national and global goals to eliminate NTDs and achieve universal health coverage. NTDs also disproportionately affect women and efforts to reduce the incidence of these diseases will also help to redress gender inequalities that persist in health.

### Limitations

The study was postponed by six months due to the COVID-19 pandemic and data collection methods were adapted to respect infection prevention and control measures. Focus group discussions were replaced with KIIs to limit person-to-person contact, which meant fewer participants were recruited due to time limitations, resulting in a narrower range of views and opinions being captured. The pandemic also hindered the full sample size being obtained in the initial assessments, however, this was compensated for by over-achieving the sample size in the final assessments. The research team worked hard with the district, regional and national health officials to ensure medicines and supplies were allocated to the research site, however, stock-outs were occasionally experienced. When this occurred, the team procured the necessary items as required. The comparison of CCA and microscopy to determine schistosomiasis case positivity rate pre-and post-implementation of the intervention, with the increased detection possibly due to the increased sensitivity of point-of-care CCA tests compared to microscopy, opposed to other intervention factors.

## Conclusion

The capacity of all enrolled health facilities for detecting, managing and recording target NTDs, through the introduction of the intervention, improved over time. Similarly, the use of intervention materials by health workers and HEWs increased over the intervention period. The intervention tools proved to be highly acceptable to health workers and HEWs, who viewed them as helpful, relevant and easy to use. Furthermore, patients and caregivers appeared to be satisfied with the new NTD services offered at the different levels of the health system. The intervention was also seen to improve care-seeking behaviours over time and be cost-effective.

To strengthen the detection, management and reporting of target NTDs in Ethiopia health workers and HEWs should continue to receive regular training and supportive supervision. Support to ensure sustainable, robust routine data collection for NTDs must also continue. Further work is needed to evaluate the feasibility and effectiveness of this intervention using a hybrid effectiveness-implementation evaluation design. Further work is also needed to integrate podoconiosis and lymphatic filariasis into the DHIS2 reporting system to ensure the effective management and reporting of these diseases.

## Funding

This work received financial support from the Coalition for Operational Research on Neglected Tropical Diseases, which is funded at The Task Force for Global Health primarily by the Bill & Melinda Gates Foundation, by the United States Agency for International Development through its Neglected Tropical Diseases Program, and with UK aid from the British people.

## Data Availability

All relevant data are within the manuscript or available upon request.

## Acknowledgements

We would like to thank all the health workers of Boditi primary hospital, Buge health centre and the health posts who participated in the study, along with the community members of Damot Gale woreda. Thank you to both partners of the study; the Federal Ministry of Health and the Health Bureau of the Southern Nations, Nationalities and Peoples’ Region. Authors would especially like to thank Lisa Chestnutt and Ani Steele for their support in finalising this manuscript.

## Supplementary information

**S1 – Table**

**Table 1.** Characteristics of patients enrolled in the baseline and endline assessment.

| Variable |  | Baseline | Endline |
| --- | --- | --- | --- |
| *Health facility type* | Hospital | 10 (23%) | 45 (34.9%) |
|  | Health centre | 21 (49%) | 58 (45%) |
|  | Health post | 12 (28%) | 26 (20.2%) |
| *Sex* | Female | 23 (53.5%) | 74 (57.4%) |
|  | Male | 20 (46.5%) | 55 (42%) |
| *Age category* | <5 | 4 (9.3%) | 18 (14%) |
|  | 5-15 | 2 (4.7%) | 11 (8.5%) |
|  | 15+ | 37 (86%) | 100 (77.5%) |
| Total |  | 43 | 129 |

## Notes

### Competing Interest Statement

The authors have declared no competing interest.

### Funding Statement

Yes

### Author Declarations

This study was approved in June 2020 by the SNNPR Public Health Institute Research Ethical Committee.

